# Estimating heart rate variability using facial video photoplethysmography: a pilot validation study

**DOI:** 10.1101/2025.02.13.25322028

**Authors:** Leszek Pstras, Tymoteusz Okupnik, Beata Ponikowska, Bartlomiej Paleczny

## Abstract

**Introduction:** Video photoplethysmography (vPPG) employs a digital camera to detect blood pulsations in the skin vasculature, which can be used to estimate various physiological parameters. In this study, we investigate the accuracy and precision of two heart rate variability (HRV) indices estimated using a smartphone camera and the facial vPPG technology Shen.AI Vitals.

**Methods:** The study group included 35 healthy volunteers (17 females) with median age 25 years (range 20–42 years). The subjects were in a sitting position, keeping their heads relatively still. A smartphone mounted on a tripod was used to acquire 1-min video recordings of participants’ faces. In parallel, a 1-lead chest electrocardiogram (ECG) was recorded to obtain reference values of two analysed time-domain HRV indices: SDNN and lnRMSSD.

**Results:** For SDNN, the mean absolute error (MAE) was 3.5 ms (11.0% in relative terms) and the root-mean-square error (RMSE) was 4.5 ms (15.7%). For lnRMSSD, the MAE was 0.24 (7.3%) and RMSE was 0.31 (9.9%). Correlations between the vPPG-based and ECG-based HRV values were strong, with the Pearson correlation coefficient of 0.98 for SDNN and 0.88 for lnRMSSD (P < 0.001 in both cases).

**Conclusions:** In a young, white population, the tested vPPG technology estimated HRV indices (SDNN and lnRMSSD) with acceptable accuracy in most subjects, with a slight systematic overestimation, especially for low values. The results should be confirmed in a larger study with greater diversity in age and skin tone.

## INTRODUCTION

Video photoplethysmography (vPPG), also known as remote or imaging photoplethysmography, is an optical technique of recording blood pulsations in the skin vasculature, which can be used to estimate vital signs or other physiological parameters [1, 2]. Specifically, vPPG enables recording of fluctuations in the intensity of ambient light reflected from the skin caused by changes in the amount of light absorbed by the blood in the superficial vessels due to their cyclic pulsation. These fluctuations are invisible to the naked eye but usually strong enough to be detected by a digital camera. vPPG provides therefore the possibility of recording PPG signals in a contactless manner without the need of special light sources and using a camera as a light sensor. This makes it possible to use cameras built into mobile devices, such as smartphones, tablets, or laptops, to measure or monitor various physiological parameters [3, 4].

Similar to standard PPG signals, vPPG signals can be used to estimate the time intervals between successive blood pulsations, which are relatively similar to the time intervals between heartbeats (although not exactly the same [5, 6]). These time intervals can be subsequently used to calculate (estimate) heart rate (HR) as well as various indices of heart rate variability (HRV) describing the variability of the time intervals between successive heartbeats [7]. HRV is a well-known marker of autonomic nervous system activity with various applications [8, 9]. A reduced HRV indicates a less adaptive autonomic regulation of the heart (i.e. an impaired ability to respond to various stimuli) and is an independent risk factor for cardiovascular events and mortality [10, 11]. Low HRV is also associated with higher risk for developing hypertension [12, 13] or cardiovascular disease [14]. HRV can also be used to predict the risk of various adverse events, for example the risk of myocardial ischemia in patients without known coronary artery disease [15], the risk of renal function deterioration in patients with chronic kidney disease [16], or the risk of falls [17]. Beyond clinical applications, HRV can be used to monitor and manage the training and recovery process in athletes [18–20], particularly in endurance sports, such as long-distance running, swimming, cycling, rowing, or cross-country skiing [19], but also in other sports [20–22].

In this study, we investigated HRV indices estimated using facial vPPG technology called Shen.AI Vitals developed by MX Labs (Tallinn, Estonia). This technology employs mobile device cameras to acquire 1-min vPPG signals from several regions of the face (by detecting pulsatile changes in the intensity of ambient light reflected from the facial skin) and then analyses these signals to estimate various physiological parameters, including the HRV index -– SDNN, i.e. the standard deviation of the time intervals between successive normal heartbeats (default), or lnRMSSD, i.e. the natural logarithm of the root mean square of successive differences in time intervals between heartbeats (optional).

SDNN is the most common HRV index in the time domain [7, 8, 23] and captures the total variability in inter-beat time intervals in the given time period (quantified by their standard deviation), thus providing an overall measure of current cardiac and autonomic nervous system activity [23, 24]. In turn, RMSSD quantifies the variability of successive inter-beat time intervals, thus focusing on high-frequency HR oscillations, reflecting mostly (albeit not exclusively) the activity of the parasympathetic nervous system [25]. Taking the logarithm of HRV indices is a common method to achieve a more normal distribution [7]. In particular, the natural logarithm of RMSSD (lnRMSSD) is often used to monitor the vagal-related adaptation to physical training and post-training recovery [18, 19, 26].

The aim of our study was to investigate the accuracy and precision of SDNN and lnRMSSD values estimated by the Shen.AI Vitals algorithms based on vPPG signals recorded with a smartphone camera, as compared with reference values obtained from simultaneously-recorded electrocardiograms (ECG).

## METHODS

### Subjects

We recruited 38 healthy volunteers (median age 25 years), including 20 females. Written informed consents have been obtained from all participants, and the study was approved by the Bioethics Committee of the Wroclaw Medical University (approval number 227/2022). Data from three subjects had to be excluded due to excessive noise or artefacts in the ECG signals; therefore, the final analysis was based on data from 35 subjects (17 females). For more details on the study participants, see Table 1.

**Table 1.** Characteristics of the 35 study participants. Data presented as median [range].

|  |  |
| --- | --- |
| Age, years | 25 [20 – 42] |
| Weight, kg | 64 [47 – 104] |
| Height, cm | 174 [160 – 193] |
| Systolic blood pressure<sup>1</sup>, mmHg | 115 [100 – 140] |
| Diastolic blood pressure<sup>1</sup>, mmHg | 80 [68 – 94] |

<sup>1</sup> average result of two upper arm cuff BP measurements (before and after video recording) after one discarded measurement

### Study overview

Video recordings were taken with the front camera of a smartphone mounted on a tripod at a distance of around 50 cm from participant’s face, at a height adjusted to the given participant (see Fig. 1). The subjects were asked to keep their heads steady during the video recording, to breathe normally, and to refrain from speaking. One measurement per participant was performed under resting conditions (two other measurements followed in other conditions but were not analysed in the present study). The study took place in a laboratory room with the window blinds closed and the ceiling lights on. To ensure sufficient light levels, a ring-shaped LED lamp (with a natural light colour) was mounted on the tripod behind the smartphone. The subjects did not engage in any physical activity before participating in the study. During the study, they were sitting with their back supported. Video recordings were initiated around 5 min after connecting all data acquisition devices.

**Fig. 1.**
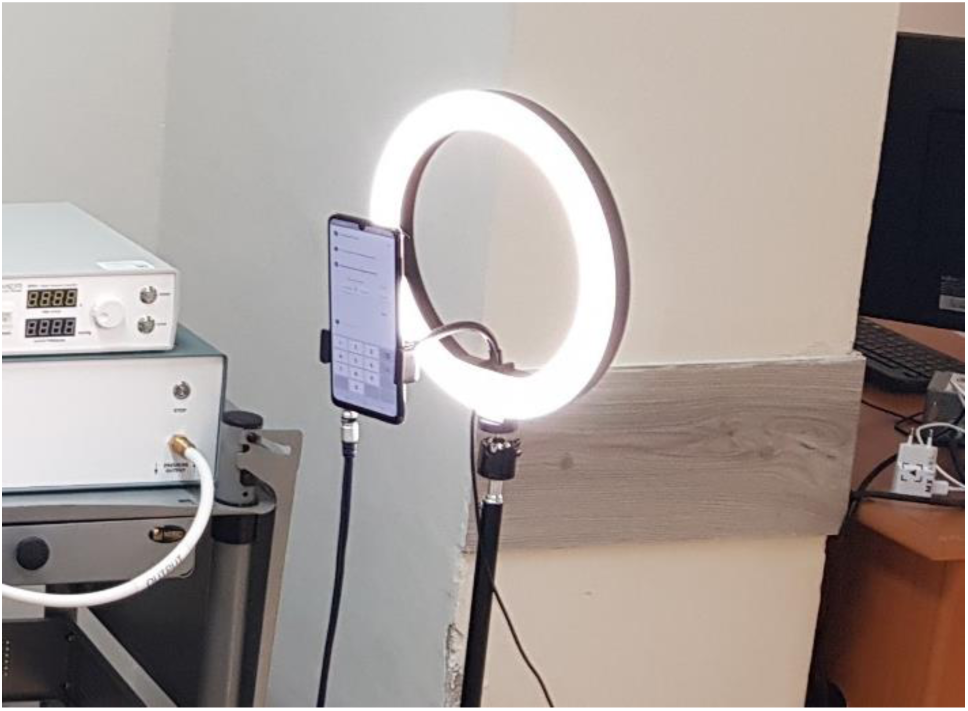
Measurement setup. The phone and the auxiliary LED lamp were mounted on a tripod at a height adjusted to the given participant sitting approximately 50 cm from the phone.

### Equipment

The front camera of the smartphone (Samsung Galaxy A22) was used for video recordings with the following parameters: video resolution 1080p, f/2.2, frame rate 30 fps. 1-lead chest ECG was recorded simultaneously in Einthoven Lead I configuration (Bio Amp ML132 module and the PowerLab data acquisition system, ADInstruments, Dunedin, New Zealand). Blood pressure was measured oscillometrically using a brachial cuff (Omron M4 Intelli IT). Other measurements (not analysed in the present study) included continuous monitoring of blood pressure (with a finger cuff), breathing rate (using a chest belt), and blood oxygen saturation (with an ear pulse oximeter).

### Video collection

The vPPG mobile app developed by MX Labs (called Heart Monitor) performs a 1-min video recording of the face, which is analysed using the computing power of the mobile device to estimate various physiological parameters displayed to the user during or after the video measurement. The present study was conducted using a special, research mobile app (provided to us by MX Labs), which records a facial video that is subsequently sent to a cloud server to be analysed at a later time using the Shen.AI Vitals algorithms. This was done for two reasons. First, this research app included a special sound signal sent by the app to the PowerLab system at the beginning and end of each video recording to enable synchronization of the ECG signal with the video, which was crucial for our study. Second, using this approach, we were blinded to the values of HRV indices estimated by the tested technology. These values were provided to us by MX Labs only after the study was completed, so that we could compare them with reference values that we had independently calculated from ECG. Since this research app recorded 2-min videos, only the first minute of each recording was analysed in the present study.

### ECG processing

ECG signals were exported to LabChart 8 (ADInstruments) for automatic detection of QRS complexes and determination of the time intervals between successive R peaks (in a few cases, R peaks were marked manually). Data from three subjects had to be excluded due to high levels of noise or artefacts in the ECG signals. The R-R intervals (in ms) and their time stamps were exported to a text file, which was subsequently processed using MATLAB (The Mathworks Inc., USA). First, all R-R intervals contained within the first minute of the video recording were selected. Second, any R-R intervals that were misidentified due to falsely detected R-peaks were corrected by merging two adjacent R-R intervals if their total length was between 80% and 120% of the median length of all identified R-R intervals.

To allow for a fair comparison, for the calculation of SDNN, similarly as done in the tested vPPG technology, we removed possible low-frequency trends from the R-R interval time series (R-R tachogram), so that the calculated SDNN value was not affected by possible larger changes in HR during the measurement (note that even though all measurements were taken under resting conditions and the subjects were seated for a few minutes before the measurement, some slow changes in HR during the measurement were still possible and were indeed observed in some of our subjects). To this end, we first interpolated the R-R tachograms to 4 Hz using a piecewise cubic hermite interpolating polynomial (PCHIP) [27], then we applied the detrending technique proposed by Tarvainen et al. [28] with lambda = 500, which corresponds to the cut-off frequency of 0.035 Hz [29], and finally we interpolated the detrended time series back to the original time points (i.e. the end times of the original R-R intervals) to obtain the final detrended tachogram of R-R intervals (in ms, rounded to the whole number).

### Calculation of reference HRV values

SDNN was calculated as the population standard deviation of all R-R intervals in the analysed 1-min period (from the detrended R-R tachogram). Given the manual and/or automatic correction of falsely detected R-peaks, the analysed intervals were treated as corresponding to normal heartbeats. The second HRV index, i.e. lnRMSSD, was calculated as the natural logarithm of the root mean square of successive differences in R-R intervals in the analysed 1-min period (here, any potential low-frequency trends would have none or minimal impact on the results, and hence we analysed the original, non-detrended R-R tachogram in order not to introduce any potential errors in the process of signal interpolation). All reference HRV values were rounded similarly as the tested values, i.e. to the nearest whole number for SDNN (in ms) and to one tenth for lnRMSSD (dimensionless).

### Statistics

We compared the values of HRV indices estimated by the tested vPPG technology with the reference ECG-based values using mean errors (ME), standard deviations of errors (SD), mean absolute errors (MAE), and root-mean-square errors (RMSE) in both absolute terms and relative terms, i.e. with respect to reference values. Correlation between the tested and reference values was assessed using the Pearson correlation coefficient (r). Statistical significance was set at P = 0.05. The agreement between the tested and reference values was visualized using scatter plots.

## RESULTS

Table 2 presents a summary of HR values and the values of the two analysed HRV indices (SDNN and lnRMSSD) determined in the study participants based on ECG recordings. In Table 3 we show various measures of accuracy and precision of the two HRV indices estimated using the tested vPPG technology (compared to ECG-based values), in both absolute and relative terms, i.e. with respect to reference values.

**Table 2.**
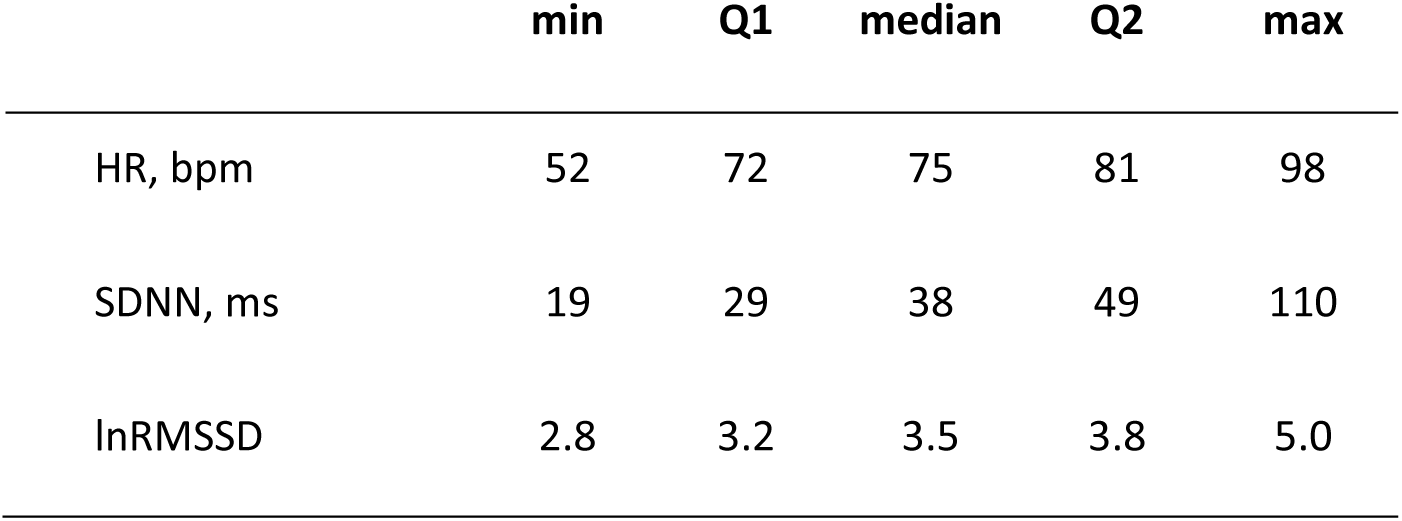
Summary of ECG-based values of average heart rate (HR) and 1-min heart rate variability indices across all study participants (n=35). SDNN – standard deviation of normal heartbeat time intervals, lnRMSSD – natural logarithm of the root mean square of successive differences in time intervals between heartbeats.

|  | **min** | **Q1** | **median** | **Q2** | **max** |
| --- | --- | --- | --- | --- | --- |
| HR, bpm | 52 | 72 | 75 | 81 | 98 |
| SDNN, ms | 19 | 29 | 38 | 49 | 110 |
| lnRMSSD | 2.8 | 3.2 | 3.5 | 3.8 | 5.0 |

**Table 3.** Measures of accuracy and precision of two heart rate variability indices estimated using the Shen.AI Vitals technology as compared with ECG-based values, expressed in absolute terms and in relative terms (as percentage of ECG-based values). Abbreviations: SDNN – standard deviation of normal heartbeat time intervals, lnRMSSD – natural logarithm of the root mean square of successive differences in time intervals between heartbeats, ME – mean error, SD – standard deviation of errors, MAE – mean absolute error, RMSE – root-mean-square error, MPE – mean percentage error, SDPE – standard deviation of percentage errors, MAPE – mean absolute percentage error, RMSPE – root-mean-square percentage error.

|  | *Absolute errors* |  |  |  | *Relative errors* |  |  |  |
| --- | --- | --- | --- | --- | --- | --- | --- | --- |
|  | **ME** | **SD** | **MAE** | **RMSE** | **MPE** | **SDPE** | **MAPE** | **RMSPE** |
| SDNN, ms | 2.80 | 3.62 | 3.54 | 4.54 | 9.59% | 12.60% | 10.95% | 15.69% |
| lnRMSSD | 0.23 | 0.21 | 0.24 | 0.31 | 7.17% | 6.93% | 7.28% | 9.90% |

Fig. 2 presents the vPPG-based HRV indices plotted against the values obtained from ECG. For both SDNN and lnRMSSD, the correlation between the values obtained by the two methods was very strong (r = 0.98 for SDNN and r = 0.88 for lnRMSSD, P < 0.001 in both cases).

**Fig. 2.**
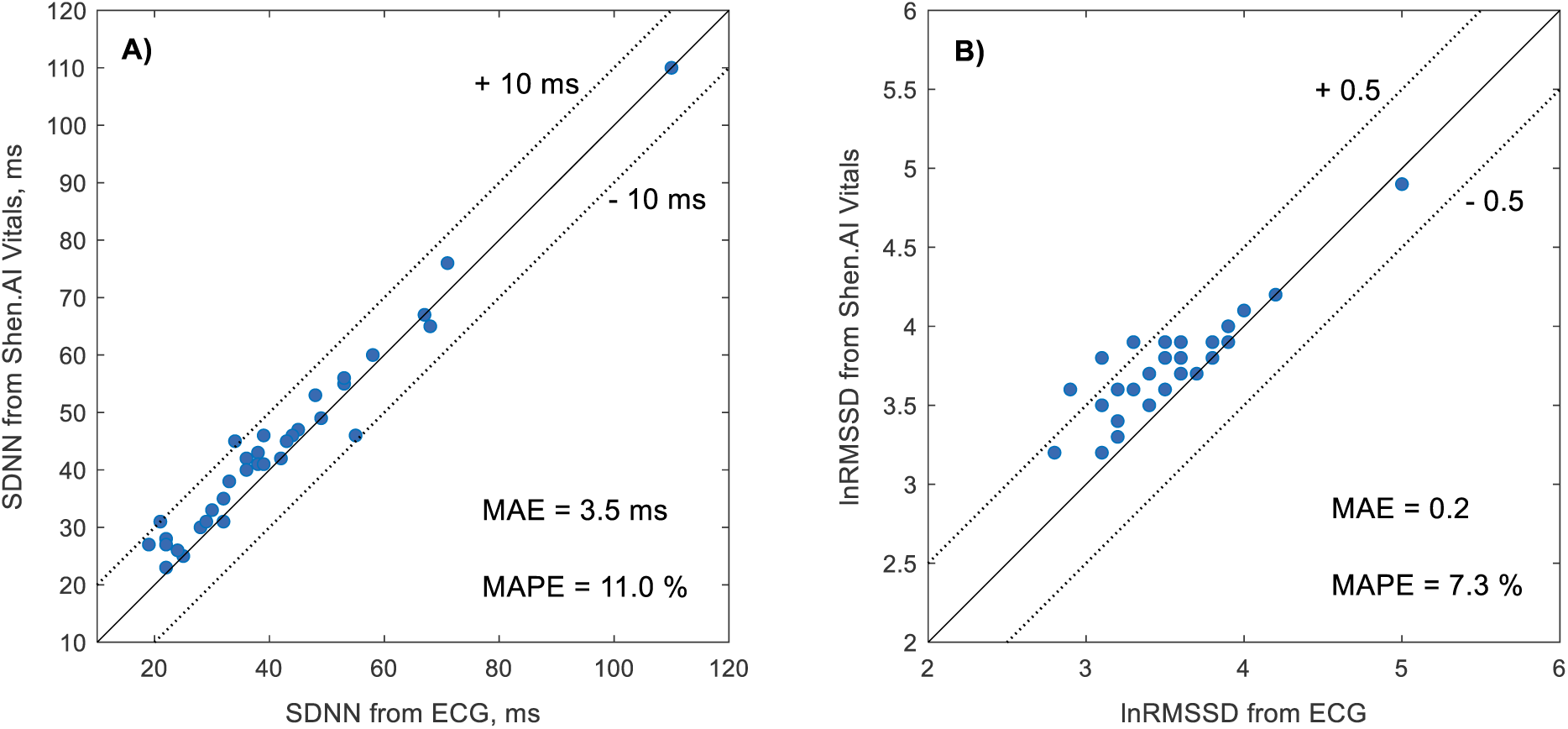
Heart rate variability indices estimated by the Shen.AI Vitals technology during 1-min facial video measurements plotted against reference values obtained from simultaneously recorded electrocardiogram (ECG). **A)** Standard deviation of the time intervals between normal heartbeats (SDNN). **B)** Natural logarithm of the root mean square of successive differences in time intervals between heartbeats (lnRMSSD). Dotted lines indicate arbitrarily chosen error limits. Abbreviations: MAE – mean absolute error, MAPE – mean absolute percentage error.

In almost all cases (34 out of 35), SDNN values estimated by the tested technology were within 10 ms of the ECG-based reference values (the remaining case had an error of 11 ms). For lnRMSSD, almost 90% of vPPG-based values (31 out of 35) were within 0.5 of the reference values.

For SDNN, the mean error between the vPPG-based and ECG-based values was 2.8 ms (9.6% in relative terms), suggesting a slight systematic overestimation of SDNN by the tested technology, particularly for low SDNN values. Similarly, we observed overestimation of lnRMSSD values by around 0.2 (7.2% in relative terms), especially for low values.

In the vast majority of cases, absolute percentage errors (APE) did not exceed 10% for both SDNN and lnRMSSD (see Table 4 and Fig. 3). Mean absolute percentage errors (MAPE) were 11.0% and 7.3% for SDNN and lnRMSSD, respectively. In the case of lnRMSSD, errors never exceeded 25%, whereas in the case of SDNN, this was true for 31 subjects (89%) – see Table 4.

**Fig. 3.**
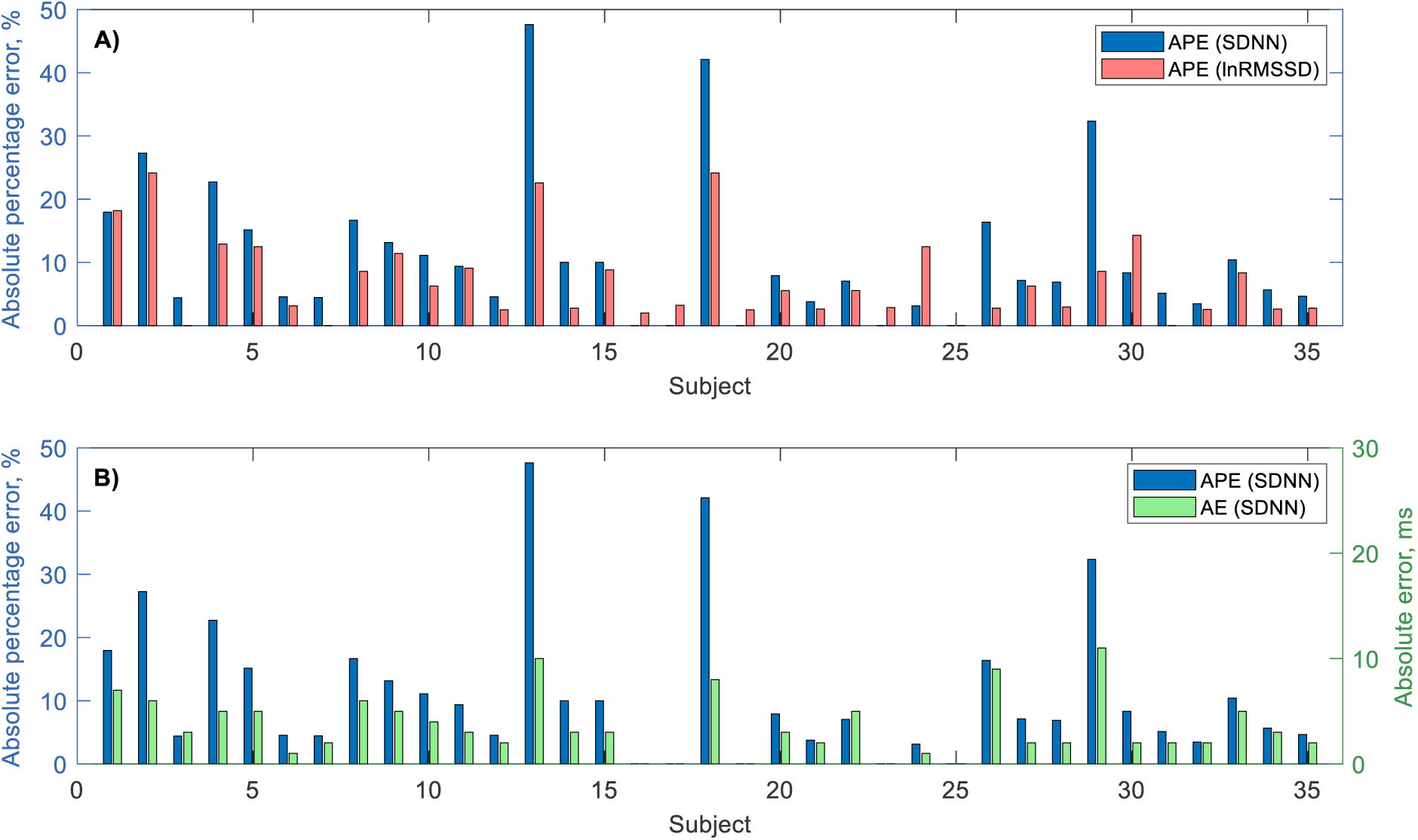
Errors in heart rate variability indices estimated by the Shen.AI Vitals technology from 1-min facial video recordings in 35 subjects (as compared with ECG-based values). **A)** Absolute percentage errors (APE) in SDNN (blue bar, left) and lnRMSSD (red bar, right). **B)** Absolute percentage errors (APE) in SDNN (blue bar, left) and absolute errors (AE) in SDNN (green bar, right). Abbreviations: SDNN – standard deviation of normal heartbeat time intervals, lnRMSSD – natural logarithm of the root mean square of successive differences in time intervals between heartbeats.

**Table 4.** Distribution of mean absolute percentage errors (MAPE) in SDNN and lnRMSSD values estimated by the Shen.AI Vitals technology in 35 studied subjects as compared to reference values obtain from simultaneously recorded electrocardiogram.

| MAPE | SDNN | lnRMSSD |
| --- | --- | --- |
| ≤ 5 % | 13 (37.1%) | 17 (48.6%) |
| ≤ 10 % | 23 (65.7%) | 26 (74.3%) |
| ≤ 15 % | 26 (74.3%) | 31 (88.6%) |
| ≤ 20 % | 30 (85.7%) | 32 (91.4%) |
| ≤ 25 % | 31 (88.6%) | 35 (100.0%) |

## DISCUSSION

In this study, we investigated the accuracy and precision of two popular time-domain HRV indices – SDNN and lnRMSSD – estimated by the tested vPPG technology based on a 1-min facial video taken with a smartphone camera. HRV indices have been traditionally determined based on 5-min measurements. However, shorter measurements, referred to as ultra short-term HRV, are becoming more and more common [30, 31]. For RMSSD, it has been shown that even measurements shorter than 60 s should be sufficient to obtain results similar to standard 5-min measurements [32–34], whereas for SDNN the recommendations vary from 30 s [32], through 60 s [35], up to 4 min [33].

### Our results

The correlation between the vPPG-based and ECG-based SDNN values was very high (r = 0.98) and the mean absolute error (MAE) was relatively low (3.5 ms); however, the mean absolute percentage error (MAPE) was slightly above 10% (11.0%). For lnRMSSD, the correlation with ECG-based values was somewhat lower (r = 0.88), which is mainly due to relatively lower range of the observed values, with MAE of 0.2 and MAPE below 10% (7.3%). For SDNN, in a few subjects we observed relatively high percentage errors, exceeding 40% in two cases. However, in these cases, SDNN was relatively low, and hence in absolute terms these errors did not exceed 10 ms (see Fig. 3). For lnRMSSD, the relative accuracy was better, as the errors never exceeded 25%.

### Previous studies

Similar results for SDNN (r = 0.98, MAE = 3.5 ms) were found in a study of 15 subjects, in whom vPPG-based values were compared to values obtained from classic finger PPG [36]. A slightly higher MAE (4.9 ms) was reported in a study on 14 subjects (with classic PPG as reference) [37], while a slightly lower MAE (2.7 ms) was reported in another vPPG study on 5 subjects (with chest-band ECG as reference) [38]. Much higher values of MAE (8.1 ms [39] and 18.0 ms [40]) were reported in studies based on a publicly available dataset with 59 video recordings of 10 subjects (with finger PPG as reference), although that dataset includes also cases where participants were asked to talk or move their head during the recordings [41]. In another study that used several publicly available datasets, MAE for SDNN was reported to range from 6.2 to 17.5 ms [42]. Note, however, that the results of different vPPG studies should be compared with great caution, not only because of possible differences in the behaviour and stability of the subjects, but also because of probable differences in the characteristics of the studied subjects (in terms of age, skin tone, or possible makeup), as well as because of the possible differences in terms of lighting, recording equipment, or the measurement setup (e.g. body position or distance from the camera). Also, the length of the measurement (video recording) may greatly influence the agreement between vPPG-based and reference HRV values.

In particular, the agreement for SDNN is expected to increase with the recording length [39], because the longer the measurement, the higher the SDNN [33], and for higher values the errors are typically lower. For instance, a lower MAE for SDNN (2.8 ms) was reported in a study on 55 subjects involving 5-min video recordings with average SDNN of 48 ms (vs 41 ms in our study) [43]. On the other hand, in another study with 5-min video recordings in 8 subjects, errors in SDNN were higher than in our study (mean error 6.7 ms), despite the similar average level of SDNN (41 ms) [44].

For lnRMSSD, we have not found any studies reporting the accuracy of vPPG-based measurements; however, there have been several studies investigating the accuracy of lnRMSSD from contact-based PPG measurements, which in theory are expected to be more reliable. For instance, a higher correlation of finger infrared PPG-based lnRMSSD with ECG-based values (r = 0.98) and a lower mean error (around 0.05) was reported in a study on 30 subjects [45], although in that study the average value of lnRMSSD was higher than in our study (3.9 vs 3.5), which, again, could have affected the level of accuracy. In another study on 31 subjects with finger camera-based PPG measurements, the mean error in lnRMSSD (as compared to ECG-based values) was similar to that in our study (0.2-0.3 in four different sessions), but the correlation coefficient was much lower (r = 0.59-0.76), which could have been due to the fact that the participants were not exclusively white [46]. In a study on 39 subjects investigating wrist-based PPG measurements of lnRMSSD (vs chest-band ECG), MAE was similar to that in our study (around 0.2), but MAPE was somewhat lower (5.6%) [47], although they studied 5-min measurements and the lnRMSSD values were, again, higher than in our study (likely due to supine position [48]).

### Overestimation of HRV indices

For both SDNN and lnRMSSD we observed a positive bias, i.e. overestimation of HRV indices by the tested technology, particularly for low values. These systematic differences between the vPPG-based and ECG-based HRV indices may be explained by the fact that the peaks in the vPPG signals are generally more difficult to identify compared to R peaks in ECG [49]. This is partly because the peaks in blood pulsations are generally less sharp compared to R peaks and partly because the vPPG signals are typically recorded with much more noise and artefacts compared to ECG and at a much lower frequency, although it has been suggested that the frequency of 25 Hz should be sufficient for the estimation of the most common HRV indices [50] (in our study, the vPPG signals were obtained at 30 Hz). Any errors in detecting peaks in blood pulsations translate into errors in the estimated time intervals between successive blood pulsations. This inevitably leads to underestimating the length of some interbeat time intervals and overestimating the length of other time intervals, which in consequence leads to overestimation of HRV. The fact that we observed greater overestimation for low HRV values is expected, given that at low HRV the time intervals between successive blood pulsations are generally more similar to each other (compared to the case with high HRV), and hence any error in estimating individual interbeat time intervals introduces a relatively higher error in the given HRV index. Similar overestimation of HRV by vPPG, particularly for low values, has been reported in the literature [36, 44, 45, 47].

### PRV vs HRV

It should be noted that the time intervals analysed by vPPG technology are the time intervals between successive blood pulsations in the skin vasculature, which are not exactly the same as the time intervals between the corresponding R peaks in the ECG signal. Therefore, vPPG technology measures in fact not the heart rate variability but the pulse rate variability (PRV). It has been shown in several studies that HRV and PRV indices are not necessarily equivalent [5, 6], which can be explained by the fact that PRV depends not only on HRV but also on the variability of stroke volume, blood pressure, and the speed of pulse wave propagation [51]. It has been even suggested that PRV should be treated separately from HRV [52]. However, since the tested technology reports PRV as HRV, we compared the tested values to ECG-based HRV values, thus effectively assessing the accuracy of the tested technology in estimating HRV. Besides, it has been shown that in young and healthy people in resting conditions (as investigated in our study), PRV is generally a good substitute of HRV [49, 53–57].

### Limitations

There are some limitations of our study that should be noted. First, we studied a relatively small cohort, which included predominantly young subjects. Although we expect no significant differences in terms of accuracy of vPPG-based HR estimation in older subjects, the age factor may be important for the accuracy of estimating HRV indices. It is well known that HRV decreases with age [58–61], and hence, given the relatively higher bias of vPPG technology for low HRV values, the level of accuracy of HRV estimation in older subjects could be accordingly lower. Second, we studied only white subjects, in whom vPPG technology is known to perform better compared to dark-skinned individuals [62, 63]. Finally, our study was conducted in a laboratory setting, with controlled lighting, the smartphone mounted on a tripod at participants’ head level, and the participants were required to keep their heads in a stable position and refrain from speaking or changing facial expression. Therefore, our results are not necessarily generalizable to other, more natural settings, with potentially lower facial illumination levels or a less stable head position.

## CONCLUSIONS

In young, white subjects in the sitting position, 1-min SDNN and lnRMSSD values estimated using the tested facial vPPG technology showed acceptable accuracy in most subjects (as compared with ECG-based values), with mean absolute percentage errors of 11.0% and 7.3%, respectively, and a slight systematic overestimation, especially for low HRV values. These results should be confirmed in a larger study with greater diversity in terms of participants’ age and skin tone.

## Data Availability

Anonymized data on the heart rate variability indices analyzed in the present study are available upon reasonable request to the corresponding author.

## Acknowledgements

MX Labs has provided us with a special, research version of its mobile application as well as the smartphone and the tripod with an LED lamp used in the study.

## Author contributions

L. Pstras: Conceptualization, Methodology, Software, Data curation, Formal Analysis, Writing – original draft, Writing – review & editing, Visualization.

T. Okupnik: Investigation, Data curation.

B. Ponikowska: Resources, Supervision.

B. Paleczny: Conceptualization, Supervision, Writing – review & editing.

